# First successful transplant of human immature testicular tissue after gonadotoxic therapy during childhood: complete spermatogenesis in intra-testicular grafts

**DOI:** 10.64898/2026.03.04.26347483

**Authors:** E. Goossens, V. Vloeberghs, E. De Beer, E. Delgouffe, I. Mateizel, C. Ernst, W. Waelput, I. Gies, H. Tournaye

## Abstract

**Background:** Approximately one-third of men having undergone gonadotoxic treatment in their childhood experience impaired testicular function for whom autologous transplantation of cryopreserved immature testicular tissue may represent the only opportunity to restore their fertility. Pre-clinical studies have demonstrated successful restoration of spermatogenesis following grafting of immature testicular tissue in various species, including non-human primates.

In 2002, our institution pioneered with clinical testicular tissue banking for fertility preservation in boys and adolescents. Over time, this strategy has been increasingly implemented by numerous fertility centres worldwide for patients at high risk of treatment-induced sterility.

Here, we report the first human case of autologous transplantation of frozen-thawed immature testicular tissue.

**Patient:** In 2008, testicular tissue was cryopreserved from a pre-pubertal boy diagnosed with sickle cell disease. The procedure was performed after a three-year hydroxyurea treatment and prior to receiving conditioning therapy with busulfan and cyclophosphamide for haematopoietic stem cell transplantation. One testis was surgically removed, sectioned into small fragments, and cryopreserved. Histological analysis confirmed preserved tubular architecture and the presence of spermatogonia. During the period from 2022 to 2024, the patient consistently presented with azoospermia. In December 2024, at the time of transplantation, two abnormal sperm cells were detected after enzymatic digestion.

**Method:** Eleven testicular tissue fragments (4–21 mm^3^) were thawed and autologously grafted to four intra-testicular and four subcutaneous scrotal sites. Over a one-year follow-up period, graft survival, vascularization, hormone profiles, and semen parameters were monitored. One year after transplantation, all grafts were surgically retrieved.

**Results:** Post-operative recovery was uneventful. No significant changes in endocrine or semen parameters were observed during follow-up. Whereas the intra-testicular grafts exhibited a compact parenchyma that was distinct from the looser surrounding adult parenchyma and remained readily identifiable as graft tissue, the scrotal grafts appeared more fibrotic. Enzymatic digestion of the grafts was required to recover spermatozoa, with one spermatozoon obtained from one of the four intra-testicular grafts. Histological evaluation revealed intact tubular architecture and maturation of somatic cells across all grafts. Spermatogonial stem cells, together with evidence of active spermatogenesis, were identified in two of the four intra-testicular grafts, whereas no germ cells were detected in the subcutaneous scrotal grafts.

**Conclusion:** These findings demonstrate that human immature testicular tissue can survive long-term cryostorage, revascularize after transplantation and establish spermatogenesis *in vivo*. This study provides essential proof-of-concept for fertility restoration in individuals who banked testicular tissue before puberty.

**Funding:** This study was supported by the Research Programme of FWO Vlaanderen (Research Foundation-Flanders; G0A6U25N) and VUB strategic research program (SRP89).

**Trial Registration:** NCT05414045

## INTRODUCTION

Each year, approximately 14,000 new cases of childhood and adolescent (0-19 years) cancer are diagnosed across Europe. Ongoing advances in oncological therapies have substantially improved outcome, with current survival rates reaching up to more than 80% for lymphomas, lymphoid leukaemia, nephroblastoma and retinoblastoma (Munoz Pineira *et al*., 2023). Consequently, the increasing population of long-term childhood cancer survivors highlights a clinical priority: understanding and preventing the late-onset adverse effects associated with cancer treatments. Furthermore, gonadotoxic conditioning regimens are also used for non-malignant disorders such as sickle cell disease in preparation for haematopoietic stem cell transplantation.

Boys undergoing gonadotoxic treatments face a substantial risk of sterility (Mitchell *et al*., 2022). Approximately one-third of adult men who received high-risk gonadotoxic treatment during childhood are azoospermic (Van Casteren *et al*., 2009; Borgström *et al*., 2020; Mathiesen *et al*., 2020, 2021; Kanbar *et al*., 2021; Braye *et al*., 2023; Delgouffe *et al*., 2023). Given the profound psychological and social implications of sterility, strategies for fertility preservation and restoration have gained increasing attention. As spermatogenesis begins only at puberty, pre-pubertal boys are ineligible for conventional sperm banking prior to gonadotoxic treatment. However, spermatogonial stem cells (SSCs) are present in the testes from birth. Cryopreservation of immature testicular tissue containing these SSCs followed by autologous transplantation in adulthood could become a promising strategy to avoid lifelong sterility (Tournaye *et al*., 2004; Gies *et al*., 2015). This approach was first introduced in clinical practice in 2002 at Universitair Ziekenhuis (UZ) Brussel as part of a fertility-preservation program for boys at risk of infertility (Braye *et al*., 2019).

Since then, an increasing number of fertility centres have implemented similar protocols for immature testicular tissue banking. To date, cryopreservation of immature testicular tissue has been performed in over 3,000 boys worldwide (Duffin *et al*., 2024). For these men, autologous transplantation of cryopreserved immature testicular tissue or testicular cell suspensions may be the only option for fertility restoration. However, at the moment, there is no evidence that spermatogenesis can be restored in humans through autologous transplantation.

Autologous transplantation of frozen-thawed immature testicular tissue was initially developed in murine models (Schlatt *et al*., 2002) and subsequently adapted to other species, including non-human primates. In mice, intra-testicular grafting has proven highly effective in re-establishing spermatogenesis (Van Saen *et al*., 2009), resulting in the birth of viable offspring (Shinohara *et al*., 2002). Even transplanting testicular tissue from non-human primates (Ntemou *et al*., 2019) and humans (Van Saen *et al*., 2011) to the mouse testis resulted in complete and partial restoration of spermatogenesis, respectively. In primate models, autologous grafting under the scrotal skin yielded superior outcomes compared to ectopic sites such as under the dorsal skin (Luetjens *et al*., 2008; Jahnukainen *et al*., 2012). More recently, autologous transplantation of cryopreserved pre-pubertal testicular tissue into castrated pubertal rhesus macaques led to the birth of a healthy offspring, providing compelling proof-of-concept for translational application in humans (Fayomi *et al*., 2019).

Here, we present the first documented autologous transplantation of frozen-thawed immature testicular tissue in a human subject.

## MATERIALS AND METHODS

### Ethical approval

The Ethical Review Board of UZ Brussel gave ethical approval for this work (B.U.N. 143201942579). The clinical trial is registered at clinicaltrials.gov (NCT05414045). Written informed consent for transplantation was obtained from the patient.

### Patient history

The patient, being pre-pubertal, underwent a left unilateral orchiectomy for testicular tissue banking following a three-year course of hydroxyurea therapy for sickle cell disease. This intervention preceded the initiation of myeloablative conditioning therapy with 16 mg/kg busulfan and 200 mg/kg cyclophosphamide as part of a successful curative bone marrow transplantation protocol. This equals a cyclophosphamide-equivalent dose of 5,392 mg/m^2^ (Green et al., 2014). The excised testicular tissue was mechanically fragmented, divided over twelve vials and cryopreserved using a semi-controlled protocol with 1·5 M dimethylsulphoxide (DMSO) combined with 0·15 M sucrose and 10% human serum albumin (HSA) as cryprotective medium (Baert *et al*., 2018). Three fragments were histologically analyzed. The seminiferous tubules had a normal architecture. Assessment of germ cell presence across 60 serial tissue depths demonstrated spermatogonia in only one single fragment. Even within this fragment, spermatogonia were confined to a very limited subset of seminiferous tubules, indicating clearly sparse germ cell presence (Fig. 1A).

**Figure 1.**
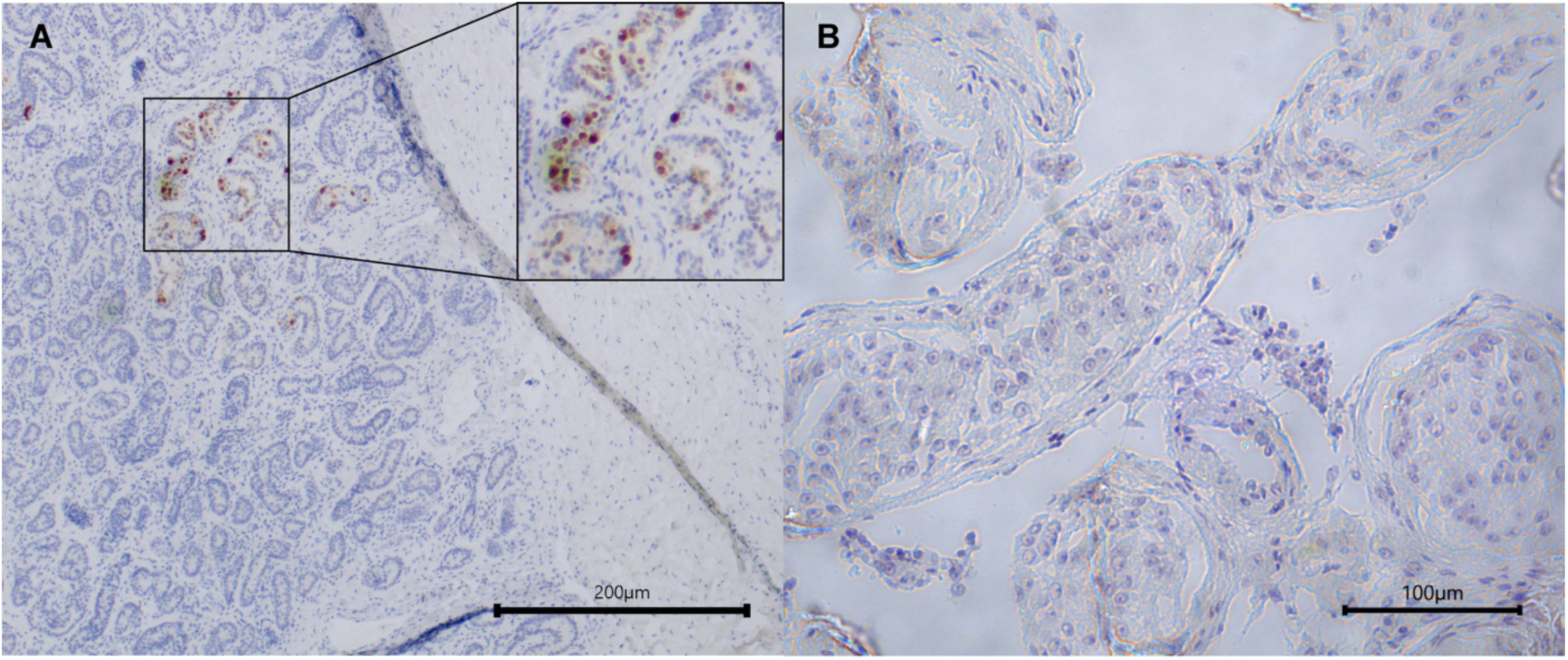
A: At the time of banking, the cryopreserved tissue showed good tubular integrity and contained (few) spermatogonia (brown MAGE-A4^+^ cells). B: At the time of transplantation, no spermatogonia were detected in the endogenous tissue.

The patient has been monitored for several years as part of a longitudinal follow-up study following gonadotoxic treatment in childhood (Delgouffe *et al*., 2023). In his early 20s, he suffered from severe obesity, with a body mass index (BMI) of >40 kg/m², hypertension (153/90 mmHg) and adipomastia. He was also diagnosed with impaired glucose tolerance and received metformin treatment (850 mg; three times daily between March 2023 and June 2023, and two times daily between June 2023 and October 2023) and endocrinological follow-up. Although gonadotrophins were high, testosterone substitution was not needed. He was advised on the importance of a healthy lifestyle and underwent gastric bypass surgery.

In 2022, he returned to Brussels IVF with a child wish. The patient was diagnosed with persistent azoospermia, as shown by three independent semen analyses conducted in March 2022, March 2024, and November 2024. He requested transplantation of the cryopreserved immature testicular tissue as part of the proof-of-concept study “Autologous testicular tissue transplantation for fertility restoration” running at UZ Brussel. At the time of screening, his weight was stable and his BMI was 28 kg/m^2^. The metformin treatment was discontinued. The patient is in a relationship and they both wish to have children in the future.

Eligibility for inclusion in the study was determined based on the following criteria: (1) a childhood diagnosis requiring sterilizing gonadotoxic therapy; (2) age ≥18 years at the time of evaluation; (3) availability of cryopreserved testicular tissue; (4) confirmed azoospermia on at least two separate assessments; (5) normal life expectancy; (6) written informed consent for the transplantation procedure, one-year post-transplant follow-up, and long-term follow-up of any offspring. Exclusion criteria included medical contra-indications to surgery and/or unstable psychological health.

All examinations and interventions took place at the Brussels IVF and the Radiology Department. Psychological support was made available for the patient throughout the clinical process.

### Screening tests

Screening tests were conducted one month before transplantation and the day of transplantation. Clinical investigation included (1) anthropometric measurements (weight, height, BMI); (2) resting blood pressure; (3) testicular volume assessment with scrotal ultrasound with a 14L5 transducer (Canon i700 Medical Systems N.V., Belgium) using the formula length x width x height x 0·71; (4) identification of parenchymal abnormalities on scrotal ultrasound; and (5) evaluation of testicular blood perfusion by colour-Doppler. Morning blood samples were collected to determine serum levels of luteinizing hormone (LH), follicle-stimulating hormone (FSH), testosterone, inhibin B (INHB) and sperm antibodies (Friberg test). Additional analyses included serological screening and standard pre-operative laboratory tests. A supplementary serum sample was cryopreserved for future research applications. A semen sample was obtained by ejaculation and analysed according to the World Health Organisation criteria (2021).

At the day of transplantation, open excisional microsurgical testicular biopsies (micro-TESE) were performed under general anaesthesia. During this procedure, 2-3 cm horizontal incisions were made to the scrotal skin, dartos muscle, and tunica vaginalis of the testis. The incision through the scrotal skin was made horizontal to stay parallel with the vascularisation of the scrotum. After opening the tunica vaginalis and exploring the testis, individual small transverse incisions in the tunica albuginea were made in four different areas of the testis (T1-T4) (Fig. 2) under assistance of an operating microscope (S7/OPMI PROergo; Zeiss, Zaventem, Belgium) under 15-20x magnification. These transverse incisions were used for sampling testicular parenchyma using microsurgical forceps. If enlarged tubules were not observed, any tubules were excised. Identifiable bleedings were coagulated. In total, four testicular biopsies were collected in individual Petri dishes containing N-2-hydroxyethylpiperazine-N-2-ethane sulfonic acid (HEPES)-buffered medium and immediately transferred to the adjacent IVF laboratory to assess the presence of spermatozoa and, consequently, suitability for cryopreservation. The testicular tissue received from the operating room was mechanically minced using Corning^®^ Cell Lifters (Corning, USA) to facilitate sperm release from the seminiferous tubules, and the resulting suspension was examined microscopically at 200x or 400x magnification. If spermatozoa were present, the suspension was centrifuged at 800 *g* for 5 min and the pellet was cryopreserved. If no spermatozoa were observed, the tissue was further processed using an enzymatic treatment to increase the likelihood of sperm retrieval (Crabbé et al., 1998; Vloeberghs et al., 2023). Briefly, the suspension was centrifuged at 1000 *g* for 5 min, and the resulted pellet was incubated in 1·5 ml ready-to-use collagenase solution (GM501 Collagenase^®^; Gynemed, Sierksdorf, Germany) for 1 h at 37° C. Following tissue digestion, the suspension was diluted with 12 ml HEPES-buffered medium, centrifuged at 100 *g* for 3 min, and the resulting supernatant was centrifuged again for 5 min at 1000 *g*. Ten microliters of the suspension was put on a slide to search for spermatozoa. If no spermatozoa were found, another 10 µl was examined (with a maximum of 5 x 10 µl). If at least one spermatozoon was observed, the testicular cell suspension was cryopreserved for possible later intracytoplasmic sperm injection (ICSI) treatment(s). For cryopreservation, the suspension was mixed 1:1 with SpermFreeze Solution (Vitrolife, Göteborg, Sweden), loaded into 0·3 ml CBS™ High Security sperm straws (Cryo Bio System, L’Aigle, France), and frozen using FREEZE CONTROL^®^ Cryopreservation system (CryoLogic, Blackburn Victoria, Australia). A separate, randomly selected testicular biopsy was submitted for histopathological evaluation. haematoxylin/periodic acid Schiff (H/PAS) staining was employed to assess the structural integrity of seminiferous tubules and the progression of spermatogenesis. Immunohistochemical staining for MAGE-A4 was conducted to identify germ cells.

**Figure 2:**
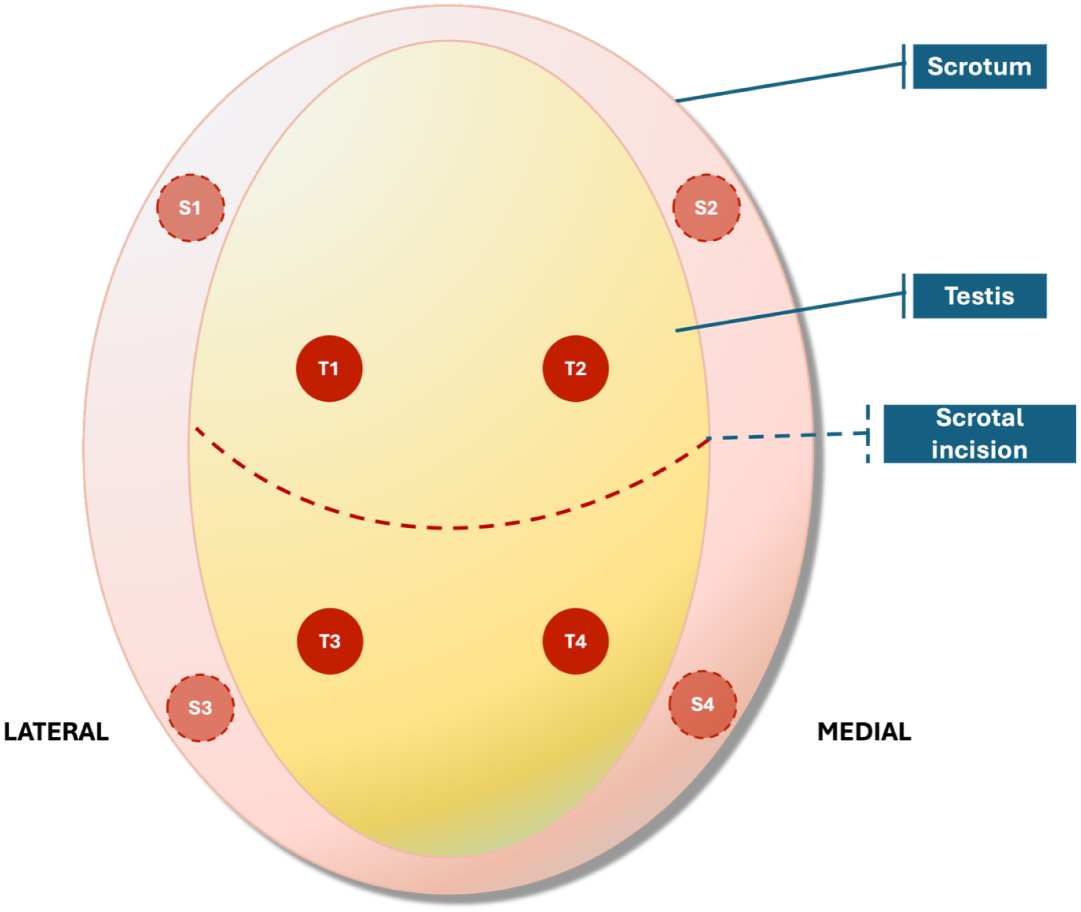
Schematic overview of the excision and transplantation sites. Testicular parenchyma was sampled from four locations (T1-T4). Eleven pre-pubertal tissue fragments were grafted into four intra-testicular locations (T1-T4) and four subcutaneous locations in the scrotum (S1-S4).

### Thawing of immature testicular tissue

Cryopreserved tissue was thawed in a 37°C water bath until the ice had melted (∼2 min), after which the tissue was transferred into Dulbecco’s Modified Eagle Medium (DMEM)/F12 supplemented with 10% HSA for 5 min on 4°C followed by exposure to DMEM/F12 for 5 min on 4°C. After thawing of the vials (n=4), the samples were transported to a clean room laboratory (type C environment), where further processing of the biopsies was carried out under a class A laminar flow hood. The biopsies were then divided into smaller fragments using a sterile scalpel. The fragments ranged in size from 0·1 cm × 0·1 cm to 0·5 cm × 0·4 cm. Upon the surgeon’s call, one fragment (or few small fragments) was individually placed in one well of a 4-well dish with DMEM/F12 and transferred immediately to the operating theatre.

At the end of the procedure, samples of freezing media and of last washing media were collected for microbiological analysis under aerobic (BD BACTEC™ Plus Aerobic/F) and anaerobic (BD BACTEC™ Plus Anaerobic/F) conditions. Culture vials were incubated in the BD BACTEC™ FX Blood Culture System for 14 days (Eur. Pharm. Monograph 2.6.27) and, in case of positive culture, the microorganisms would be identified using mass spectrometry Bruker MALDI Biotyper^®^ Sirius. Tryptic soy agar plates were used to monitor air quality and gloved fingerprints in both the clean room and laminar flow workstation.

### Transplantation procedure

Under high magnification using an operating microscope, thawed testicular tissue fragments were grafted into four distinct intra-testicular sites (T1-T4) (Fig. 2). The tissue piece(s) were tied in place with one suture through the tunica albuginea, which fixed the tissue and created small traumas in the grafted area, inducing neo-angiogenesis. Afterwards, the tunica albuginea was closed with separated stitches using non-absorbable sutures. Mono-filament polypropylene suture (Prolene 7-0) was used for graft T1 and T2, while multi-filament silk suture (Silk 8-0) was used for graft T3 and T4. The tunica vaginalis was closed in a running fashion using absorbable sutures.

Subsequently, additional thawed fragments were transplanted to four separate subcutaneous openings created in the scrotal skin (S1-S4) (Fig. 2). Anatomically, blood circulation in the scrotum originates from both lateral and medial circulation, allowing for medial and lateral grafting. To avoid compromising blood flow, the same skin incision was used to perform the testicular biopsies, to embed the testicular grafts, to create the subcutaneous pockets, and embed the subcutaneous scrotal grafts. The subcutaneous pockets were created using hydro-dissection, and haemostasis was ensured before the transplantation of the grafts. One suture with non-resorbable material was used to fixate the tissue fragment(s) in place. Once fixation was completed, the subcutaneous pocket was closed in a circular fashion using a non-absorbable suture. Each grafting site was marked in this way to ensure precise post-operative identification. Mono-filament polypropylene suture (Prolene 7-0) was used for graft S1 and S2, while multi-filament silk suture (Silk 8-0) was used for graft S3 and S4.

The dartos muscle was closed with interrupted absorbable sutures; the skin using absorbable sutures (Vicryl 3-0). Prophylactic intra-venous antibiotics (cefazoline 2 g) were administered peri-operatively to minimize the risk of infection. Throughout the whole procedure, care was taken to regularly moisten the testis with saline at 37°C to prevent tissue drying.

### Follow-up tests

The patient returned for follow-up evaluations every three months during the first year after the autologous testicular tissue grafting. Follow-up tests were identical to the screening tests. Furthermore, graft size was assessed by ultrasound and vascularization was evaluated visually using a semi-quantitative colour-Doppler vascularity score (CDVS). Vascularization was graded as follows: 0=no colour-Doppler signal; 1=minimal colour-Doppler signal; 2=moderate colour-Doppler signal; 3=marked colour-Doppler signal. In cases of very intense colour-Doppler signal a score of 4 indicating hyper-vascularity was used.

### Evaluation at one year post-transplantation

The surgery was carried out under general anaesthesia. A 2-3 cm horizontal incision was made to the scrotal skin (located on the place of the previous scar), dartos muscle, and tunica vaginalis. Grafted areas were identified by the use of non-resorbable sutures at the time of the transplantation as mentioned before. An operating microscope was used to excise the grafts. An ellipsoid-shaped incision was made around the four subcutaneous scrotal grafts. Full excision of the grafted area was followed by haemostasis. An elliptically shaped incision was made in the tunica albuginea around the marking of the four testicular grafts. The testicular parenchyma was biopsied under 15-20x magnification and the grafted area was excised, together with some endogenous testicular parenchyma surrounding the graft. Each biopsy was divided into two equal portions: one portion was processed for sperm retrieval and cryopreservation (if viable spermatozoa were present); the other portion was submitted for histological analysis to evaluate tissue integrity and cellular composition. Concurrently, a biopsy of the endogenous testicular parenchyma was obtained at a distinct site to serve as a control for the intra-testicular graft sites. In total, 12 samples were evaluated for histological analysis and for the presence of spermatozoa: i) four subcutaneous scrotal grafts, ii) four intra-testicular grafts, iii) three endogenous adult tissue adjacent to the testicular grafts (for T2, no endogenous tissue was available), and iv) one endogenous adult tissue. The dartos muscle was closed with interrupted absorbable sutures; the skin using absorbable sutures.

### Histological evaluation

The testicular specimens were fixed in alcohol formaldehyde acetic acid (AFA0060AF59001; VWR, Leuven, Belgium) for 1 h, followed by fixation in formalin overnight and embedding in paraffin. The samples were sectioned at 5 µm thickness. The overall morphology of the samples was evaluated by a haematoxylin-eosin (HE) staining. The HE staining was performed and mounted on the Histocore spectra ST (102094; Leica Biosystems, Diegem, Belgium) and Histocore Spectra CV (102095; Leica Biosystems, Diegem, Belgium). The slides were scanned using the Aperio GT450DX (93225; Leica Biosystems, Diegem, Belgium). Images were captured using the imaging software PIMSLS (version 3.1.0).

Immunohistochemical or immunofluorescence staining of the sections was performed using four markers to identify the level of germ cell differentiation within the seminiferous tubules: melanoma-associated antigen A4 (MAGE-A4), expressed by spermatogonia and primary spermatocytes; boule RNA-binding protein (BOLL), expressed in secondary spermatocytes and round spermatids; cAMP-responsive element modulator (CREM) for round spermatids; and acrosin (ACR) for acrosome visualisation in round, elongating, and elongated spermatids. Immunohistochemical staining was performed for MAGE-A4. The sections were deparaffinised in xylene and subsequently rehydrated in decreasing ethanol series (100% - 100% - 90% - 70%). After washing with phosphate buffered saline (PBS; 70011051; Life Technologies, Merelbeke, Belgium) for 5 min, endogenous peroxidase was blocked with a 0·3% peroxide solution in methanol for 30 min. After washing in PBS for 5 min, antigen retrieval was performed by incubating the slides in home-made citric acid buffer (pH 6). After blocking with normal goat serum, MAGE-A4 was applied to the sections (see Table 1 for details). The sections were incubated in a humidified chamber overnight at 4°C. PBS instead of primary antibody was applied to the negative control. The next day, after three washing steps with PBS, sections were incubated with a goat anti-mouse horse radish peroxidase (HRP)-labelled secondary antibody (Dako Real Envision Detection System; k5007; Agilent, Diegem, Belgium) at room temperature for 1 h. After washing, visualisation was performed by using 3,3ʹ-diaminobenzidine (Dako Real Envision Detection System; K500711-2, Agilent, Santa Clara, CA, USA). After counterstaining with haematoxylin, sections were subsequently dehydrated in increasing ethanol series (70% - 90% - 100% - 100%), cleared in xylene, and mounted using Prolong Gold antifade reagent (P36934; Invitrogen, Thermo Fisher Scientific, Breda, The Netherlands) for microscopic evaluation.

**Table 1.**
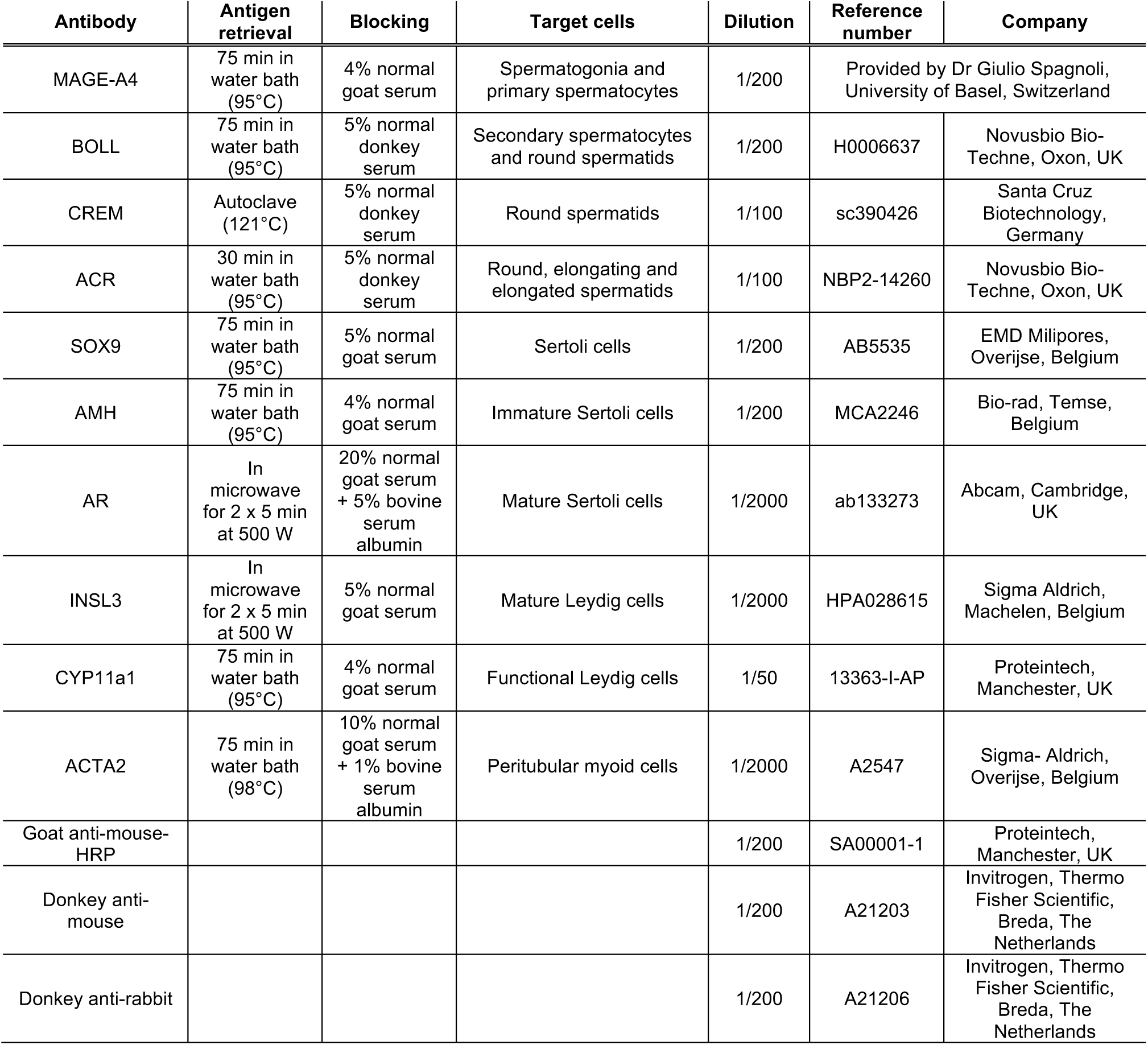
Antibody specifications.

| Antibody | Antigen retrieval | Blocking | Target cells | Dilution | Reference number | Company |
| --- | --- | --- | --- | --- | --- | --- |
| MAGE-A4 | 75 min in water bath (95°C) | 4% normal goat serum | Spermatogonia and primary spermatocytes | 1/200 | Provided by Dr Giulio Spagnoli, University of Basel, Switzerland |  |
| BOLL | 75 min in water bath (95°C) | 5% normal donkey serum | Secondary spermatocytes and round spermatids | 1/200 | H0006637 | Novusbio Bio-Techne, Oxon, UK |
| CREM | Autoclave (121°C) | 5% normal donkey serum | Round spermatids | 1/100 | sc390426 | Santa Cruz Biotechnology, Germany |
| ACR | 30 min in water bath (95°C) | 5% normal donkey serum | Round, elongating and elongated spermatids | 1/100 | NBP2-14260 | Novusbio Bio-Techne, Oxon, UK |
| SOX9 | 75 min in water bath (95°C) | 5% normal goat serum | Sertoli cells | 1/200 | AB5535 | EMD Milipores, Overijse, Belgium |
| AMH | 75 min in water bath (95°C) | 4% normal goat serum | Immature Sertoli cells | 1/200 | MCA2246 | Bio-rad, Temse, Belgium |
| AR | In microwave for 2 x 5 min at 500 W | 20% normal goat serum + 5% bovine serum albumin | Mature Sertoli cells | 1/2000 | ab133273 | Abcam, Cambridge, UK |
| INSL3 | In microwave for 2 x 5 min at 500 W | 5% normal goat serum | Mature Leydig cells | 1/2000 | HPA028615 | Sigma Aldrich, Machelen, Belgium |
| CYP11a1 | 75 min in water bath (95°C) | 4% normal goat serum | Functional Leydig cells | 1/50 | 13363-I-AP | Proteintech, Manchester, UK |
| ACTA2 | 75 min in water bath (98°C) | 10% normal goat serum + 1% bovine serum albumin | Peritubular myoid cells | 1/2000 | A2547 | Sigma- Aldrich, Overijse, Belgium |
| Goat anti-mouse-HRP |  |  |  | 1/200 | SA00001-1 | Proteintech, Manchester, UK |
| Donkey anti-mouse |  |  |  | 1/200 | A21203 | Invitrogen, Thermo Fisher Scientific, Breda, The Netherlands |
| Donkey anti-rabbit |  |  |  | 1/200 | A21206 | Invitrogen, Thermo Fisher Scientific, Breda, The Netherlands |

In the presence of MAGE-A4^+^ cells, immunofluorescence staining was performed for BOLL, CREM and ACR. After deparaffinization and rehydration, antigen retrieval was performed in home-made tris-ethylenediamine tetra-acetic acid buffer (pH 9). After blocking, primary antibodies were applied to the sections (Table 1). After an overnight incubation, the secondary antibodies donkey anti-mouse (for BOLL and CREM) or donkey anti-rabbit (for ACR) (Table 1) together with Hoechst (H3570; Life Technologies; Merelbeke, Belgium) were added to the slides and incubated for one hour. Subsequently, the slides were mounted and stored in the dark at 4°C. Histological examination was performed on an inverted microscope (IX81, Olympus, Aartselaar, Belgium). Every 20^th^ slide was analysed. Images were captured using the imaging software Toupview (version x64, 4.12.27501.20250112) at a magnification of 4x and 20x. If MAGE-A4^+^ cells were present, quantitative analysis was performed at 20x magnification. The number of MAGE-A4^+^ cells per slide was reported as mean ± standard deviation. Also, the fertility index, defined as the percentage of tubules containing complete spermatogenesis (ACR^+^ cells) was recorded.

Maturation of somatic cells was assessed as described in Delgouffe *et al*. (2025) (Table 1). Sertoli cell maturation was assessed by SOX9 (general marker), anti-Müllarian hormone (AMH, immature Sertoli cells), and androgen receptor (AR, mature Sertoli cells). The markers INSL3 (mature Leydig cells) and CYP11a1 (steroidogenesis) were used to assess the maturation status of the Leydig cells. Per sample, one slide was analysed using a confocal microscope (LSM 800, Zeiss, Zaventem, Belgium). Images were captured using the ZEN software (version 2.6, Zeiss, Zavemtem, Belgium) at a magnification of 20x.

The maturation status of the peritubular myoid cells was determined by the presence of ACTA2. Slides of three different depths were analysed using an inverted microscope (CKX5381, Olympus, Aartselaar, Belgium). Images were captured using the imaging software Toupview (version x64, 4.12.27501.20250112) at a magnification of 10x.

## RESULTS

### Screening tests

To ensure procedural efficiency and optimize patient comfort, all clinical assessments were conducted in a single day. Patient characteristics prior to transplantation and at the day of transplantation are shown in Table 2. Serology and standard pre-operative laboratory testing were normal.

**Table 2.** Patient characteristics.

|  | One month before transplantation | At day of transplantation | Follow-up visit 1 | Follow-up visit 2 | Follow-up visit 3 | Follow-up visit 4 |
| --- | --- | --- | --- | --- | --- | --- |
| Weight (kg) | 77 | 77 | 78 | 80 | 77 | 77 |
| Length (cm) | 166 | 166 | 166 | 166 | 166 | 166 |
| BMI (kg/m<sup>2</sup>) | 28 | 28 | 28 | 29 | 28 | 28 |
| Blood pressure (mmHg) | 123/75 | 99/52 | 136/93 | 122/78 | 124/83 | 137/76 |
| Volume right testis (ml) |  |  |  |  |  |  |
| Prader orchimeter | 10-11 | 11 | 11 | 11 | 11 | 12 |
| Ultrasound | 9.7 | ND | 7.5 | 7.1 | 9.0 | 8.5 |
| Testicular parenchyma | normal | normal | normal | normal | normal | normal |
| LH (IU/l) | 16.6 | 13.3 | 15.0 | 15.2 | 13.7 | 20.7 |
| FSH (IU/l) | 14.5 | 14.5 | 15.8 | 15.7 | 15.1 | 20.7 |
| Testosterone (µg/l) | 6.4 | 5.5 | 6.7 | 6.3 | 5.8 | 7.3 |
| INHB (ng/l) | 20.2 | 14.2 | 19.2 | 15.8 | 13.2 | 11.8 |
| Semen analysis | azoospermia | azoospermia | azoospermia | azoospermia | azoospermia | azoospermia |

Reference values are 1.7-8.6 IU/l for LH, 1.5-12.4 IU/l for FSH, 2.5-8.4 µg/l for testosterone and 104-439 ng/l for INHB.
Abnormal values are shown in red.
BMI: body mass index; LH: luteinising hormone; FSH: follicle stimulating hormone; INHB: inhibin B; ND: not determined.

Histological examinations were performed on a few seminiferous tubules isolated from location T1. The evaluated tubules did not contain spermatogonia (Fig. 1B). However, after enzymatic digestion of the testicular parenchyma retrieved from location T4, two immotile spermatozoa were detected. These two spermatozoa had an abnormal morphology: one had a broken flagel, the other one contained a cytoplasmic droplet and vacuoles. As per protocol, these spermatozoa were cryopreserved.

### Transplantation

Four vials containing eleven testicular tissue fragments were thawed. The size of the (combined) fragments ranged from 4 to 21 mm^3^ (Table 3). Six fragments (the three smallest ones were combined) were transplanted to four locations in the testis (Fig. 3A,B) and five fragments (the two smallest ones were combined) to four locations under the scrotal skin (Fig. 3C,D). The duration of the procedure was 2 h 53 min from the initial skin incision to the closure of the skin.

**Figure 3.**
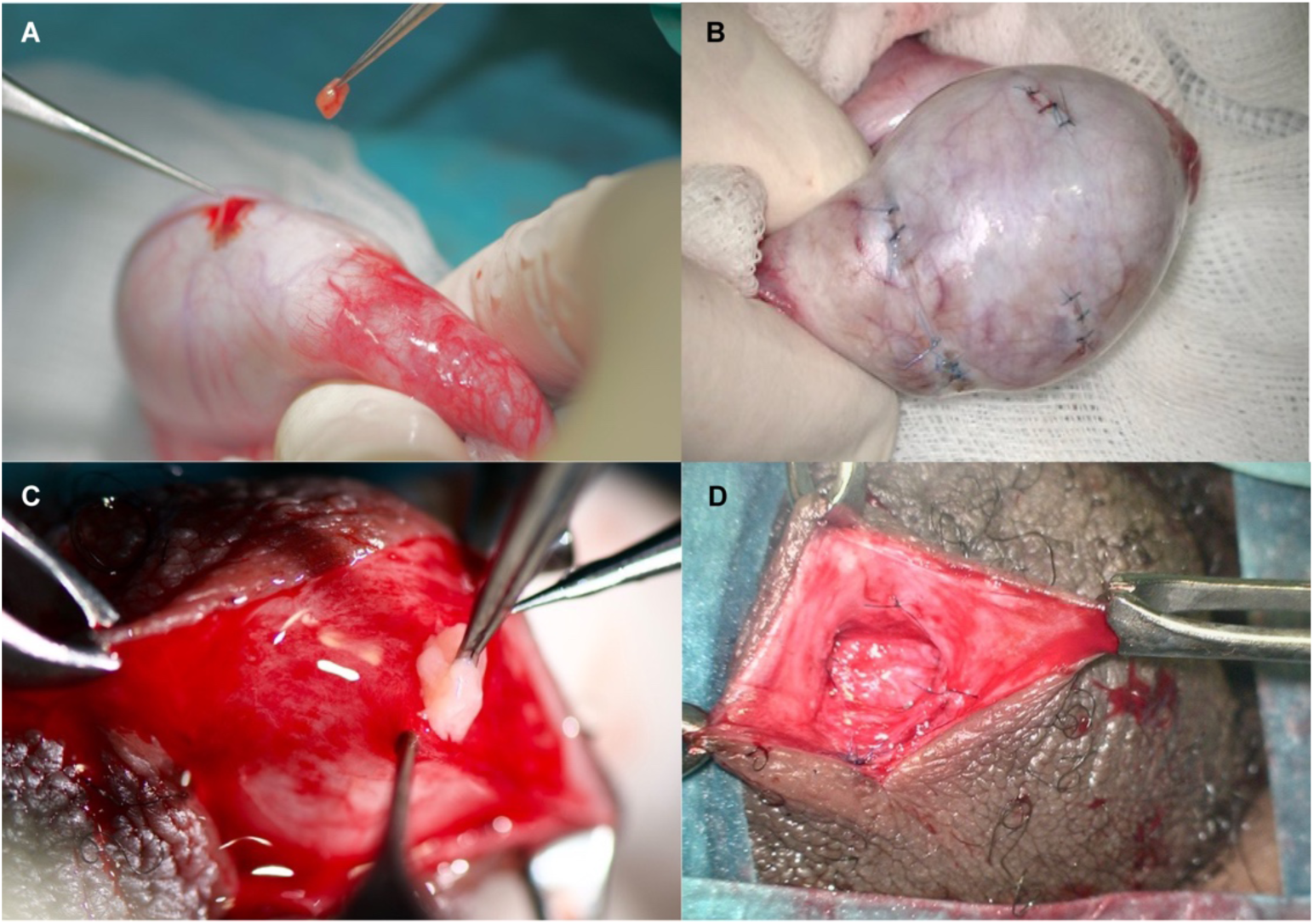
A,B: Intra-testicular grafting. C,D: Subcutaneous grafting in the scrotum.

**Table 3.**
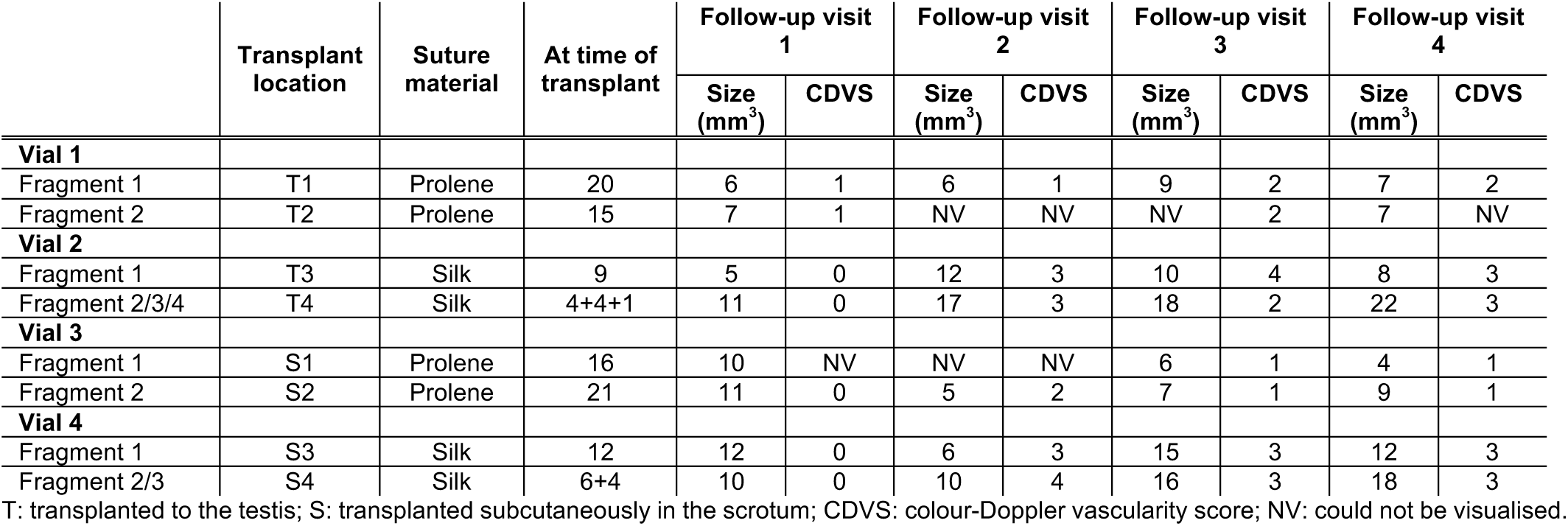
Characteristics of transplanted testicular tissue fragments on ultrasound.

All microbiological tests in the laboratory were negative, confirming that sterile conditions were maintained, and aseptic techniques were consistently applied throughout the procedure.

### Follow-up tests

Every three months, the patient returned for follow-up consultations. Blood and semen parameters as well as graft survival were evaluated (Table 2 and 3).

Gonadotrophin levels were high and INHB levels were very low. Azoospermia persisted throughout the follow-up period. Ultrasound-based follow-up assessments were conducted to evaluate graft visibility, suture integrity and vascularization. Three months post-transplantation, all grafts were clearly identifiable. A fluid collection of approximately 2 ml was present in scrotal graft S4. Six months post-transplantation, grafts T2 and S1 were not detectable. Twinkling artefacts on Doppler imaging, consistent with small calcifications, were observed in grafts T3 and S3. The previously observed fluid collection in S4 had resolved. Nine months post-transplantation, graft T2 remained undetectable, while S1 could be visualized again. Twinkling artefacts remained present in grafts T3 and S3, and now also appeared in S4. One year after grafting, all grafts were visible on ultrasound. Calcifications and twinkling artefacts remained present in grafts T3, S3 and S4. Interestingly, this time, scrotal grafts S2, S3 and S4 were palpable. Graft vascularization varied across time points and between grafts. When visible, vascularization scores ranged from 0 (no colour-Doppler signal) to 4 (hyper-vascular). Some grafts showed stable low-level vascularization, whereas others demonstrated intermittent increases in colour-Doppler signal over subsequent visits. Overall, no consistent pattern of vascularization evolution could be established across all grafts.

At all follow-up times, Prolene sutures could not be visualized by ultrasound, while silk sutures remained detectable.

### Evaluation at one year post-transplantation

At one year post-grafting, the patient was re-hospitalized for surgical removal of the grafted testicular tissue. Both Prolene and silk sutures remained identifiable, but it was easier to identify the Prolene sutures than the silk sutures. All grafts could be recovered. Both intra-testicular and subcutaneous scrotal grafts could be clearly recognized and isolated (Fig. 4). The parenchyma of the intra-testicular grafts showed better consistency than the endogenous parenchyma, which had a looser appearance. Both intra-testicular and subcutaneous scrotal grafts were significantly enlarged in size. During mincing, the scrotal grafts appeared highly fibrotic, whereas testicular grafts had a better composition and were easier to mince. Since no spermatozoa were found after mincing, minced fragments were enzymatically digested. After enzymatic digestion, one spermatozoon was found in the first 10 µl cell suspension obtained from graft T4. The remaining cell suspension was cryopreserved. In suspensions obtained from the other grafts and endogenous parenchyma samples, no (im)mature spermatozoa were detected. The remnants were collected and processed further for histological analysis.

**Figure 4.**
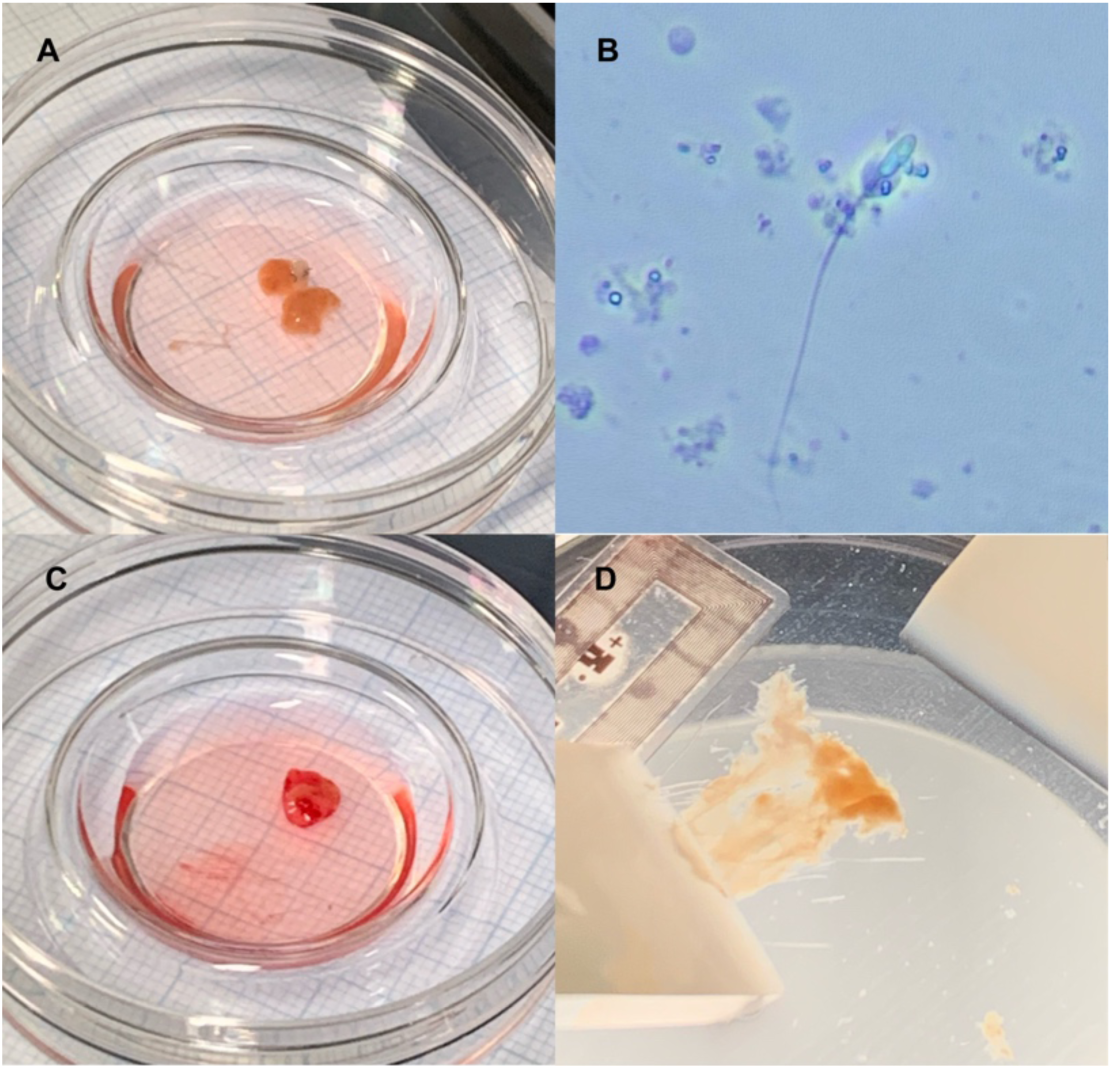
Graft appearance one year post-transplantation. A: Intra-testicular graft T1. B: Spermatozoon retrieved after enzymatic digestion of T4. C: Subcutaneous scrotal graft S4. D: Mincing of subcutaneous scrotal graft S3.

Histological evaluation of the grafts showed that tissue pieces transplanted to either the testis or subcutaneously in the scrotum survive for at least one year. In general, tubular architecture was normal. Complete spermatogenesis was observed in two out of four intra-testicular grafts (T2 and T4). Notably, in regions containing SSCs, spermatogenesis had been initiated, with differentiation progressing up to the spermatid stage (Fig. 5A,B,F,G; Fig. 6). In scrotal grafts, however, tubules contained only Sertoli cells (Fig. 5C,H). Endogenous tissue adjacent to T4 also showed complete spermatogenesis (Fig. 5D,I). Unfortunately, endogenous tissue adjacent to T2 was entirely used for searching sperm and could thus not be evaluated by histology. Other control sites in the testis did not contain germ cells (Fig. 5E,J). The histology of the remnants of the tissue that was enzymatically digested did not contain tubules, except for S1. These tubules only contained Sertoli cells (Table 4).

**Figure 5.**
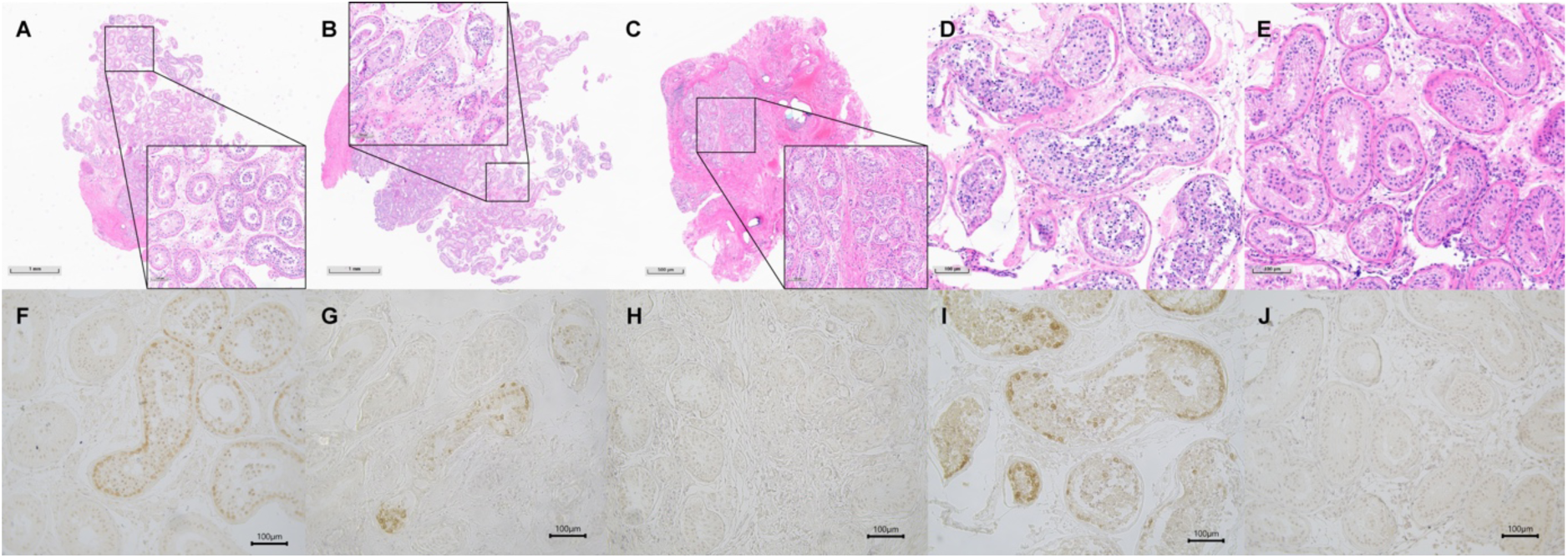
Tubular integrity and germ cell presence. A-E: haematoxylin-eosin staining. F-J: Mage-A4 staining. A,F: Spermatogenesis in intra-testicular graft T2. T2 shows one transplanted fragment. B,G: Spermatogenesis in intra-testicular graft T4. Three transplanted fragments can be recognized. C,H: Germ cell absence and extensive fibrosis in subcutaneous scrotal grafts. D,I: Endogenous tissue adjacent to graft T4 shows spermatogenesis. E,J: Control tissue from a distinct site in the testis. Tubules contain only Sertoli cells.

**Figure 6.**
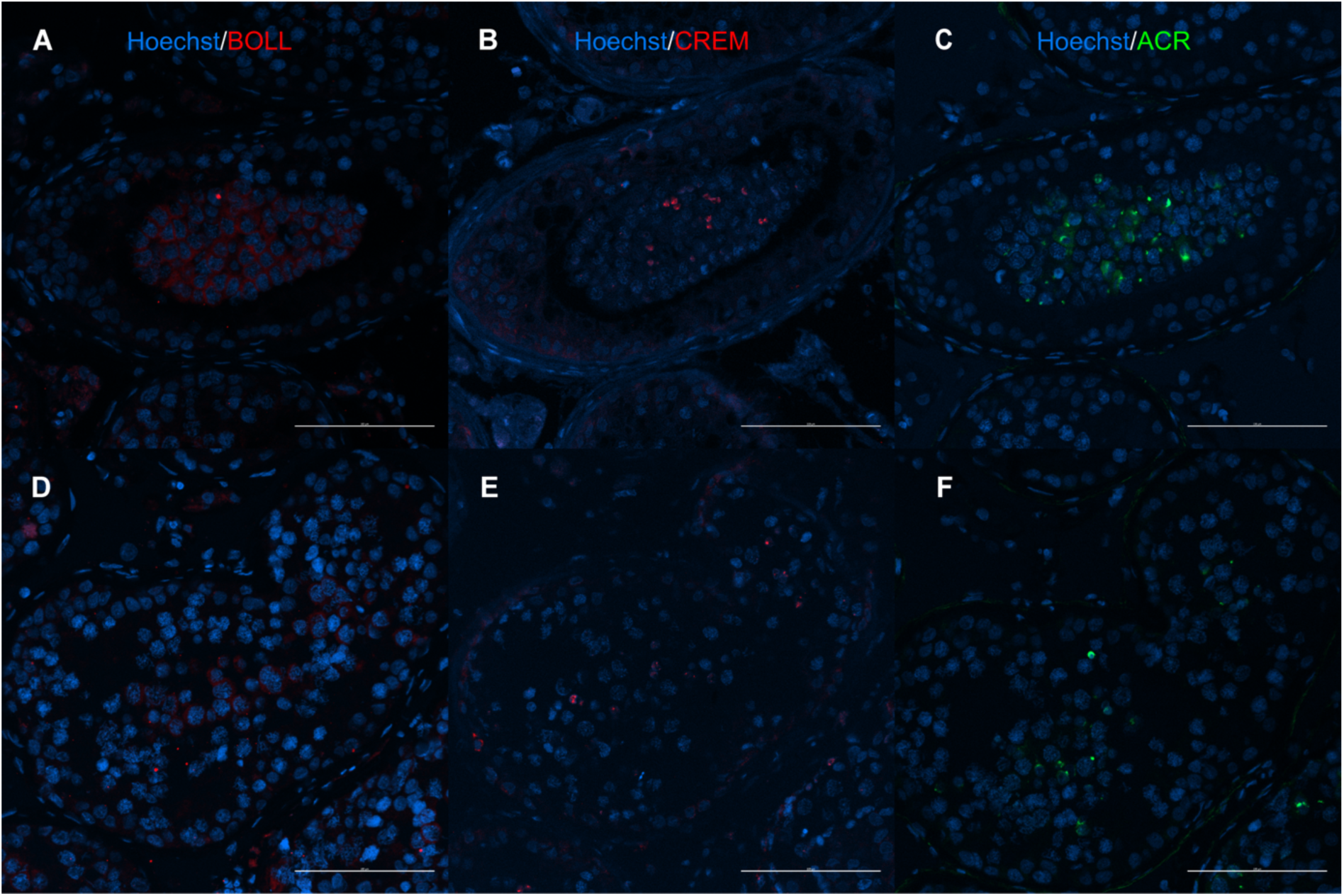
Germ cell maturation in intra-testicular grafts. A-C: Graft T2; D-E: Graft T4. A,D: Meiotic cells (BOLL^+^). B,E: Post-meiotic cells (CREM^+^). C,F: Spermatids (ACR^+^).

**Table 4.** Graft size and germ cell presence one year post-transplantation.

| Transplant location | Graft size (mm<sup>3</sup>) | Germ cell presence |  |  |  |  |  |  |  |
| --- | --- | --- | --- | --- | --- | --- | --- | --- | --- |
|  |  | Sperm by mincing | Sperm after enzymatic treatment | Spermatogenesis by HE | MAGE-A4<sup>+</sup> cells / section<sup>a</sup> | BOLL<sup>+</sup> cells | CREM<sup>+</sup> cells | ACR<sup>+</sup> cells | Fertility index<sup>a</sup> (%) |
| T1 | 51 | no | no | no | 0 | ND | ND | ND | ND |
| T2 | 85 | no | no | yes | 131 ± 71 | yes | yes | yes | 3.5 ± 1.9 |
| T3 | 45 | no | no | no | 0 | ND | ND | ND | ND |
| T4 | 110 | no | yes | yes | 89 ± 60 | yes | yes | yes | 4.7 ± 3.9 |
| S1 | 73 | no | no | no | 0 | ND | ND | ND | ND |
| S2 | 55 | no | no | no | 0 | ND | ND | ND | ND |
| S3 | 50 | no | no | no | 0 | ND | ND | ND | ND |
| S4 | 146 | no | no | no | 0 | ND | ND | ND | ND |
| TC | NA | no | no | no | 0 | ND | ND | ND | ND |
| T1<sub>end</sub> | NA | no | no | no | 0 | ND | ND | ND | ND |
| T2<sub>end</sub> | NA | no | no | ND | ND | ND | ND | ND | ND |
| T3<sub>end</sub> | NA | no | no | no | 0 | ND | ND | ND | ND |
| T4<sub>end</sub> | NA | no | no | yes | 70 ± 19 | yes | yes | yes | 4.8 ± 0.6 |

T: transplanted to the testis; S: transplanted subcutaneously in the scrotum; Tx<sub>end</sub>: endogenous tissue adjacent to the graft; TC: control tissue from a distinct site in the testis; NA: not applicable; ND: not determined; HE: haematoxylin-eosin staining; MAGE-A4: melanoma-associated antigen 4 (marker for spermatogonia and primary spermatocytes); BOLL: boule RNA binding protein (marker for secondary spermatocytes and round spermatids); CREM: cAMP responsive element modulator (marker for round spermatids); ACR: acrosin (marker for acrosome visualisation in spermatids).
<sup>a</sup>Every 300 µm, one section was counted.

Across all evaluated grafts and controls, somatic cells exhibited features of maturation, including AR^+^ Sertoli cells, INSL3^+^CYP11a1^+^ Leydig cells, and peritubular myoid cells with the expected intact ACTA staining pattern (Fig. 7-9; Table 5). Despite this, Sertoli cells continued to show weak AMH expression both in areas with and without spermatogenesis.

**Figure 7.**
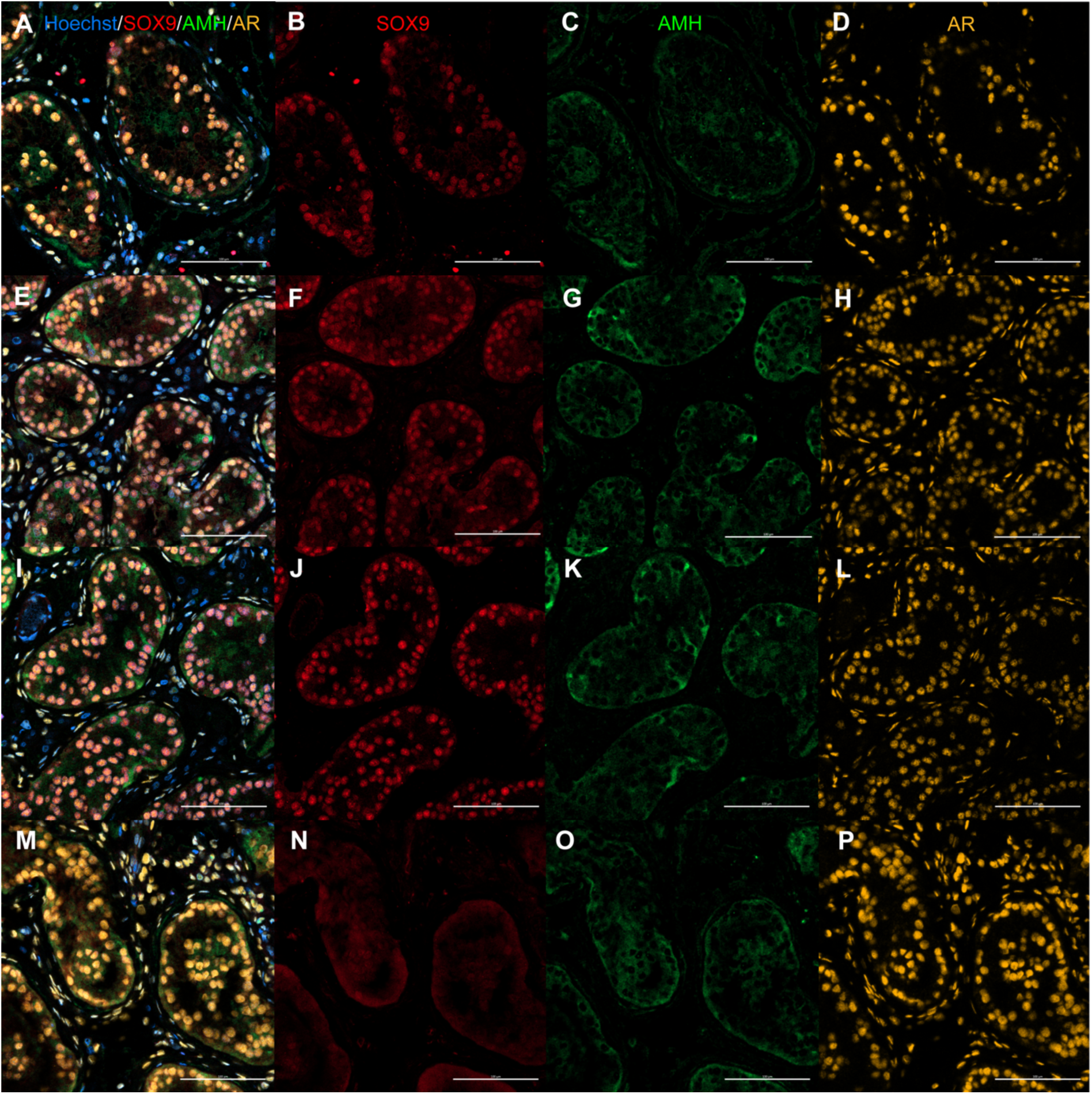
Maturation of Sertoli cells. A-D: Tubules with spermatogenesis in intra-testicular graft T4. E-H: Tubules without spermatogenesis in intra-testicular graft T4. I-L: Subcutaneous scrotal graft S2. M-P: Endogenous testicular parenchyma. Although androgen receptor (AR) was expressed in all samples, anti-Müllarian hormone (AMH) was still present. A,E,I,M: Merged pictures. B,F,J,N: SOX9 expression for Sertoli cells. C,G,K,O: AMH expression for immature Sertoli cells. D,H,L,P: AR expression for mature Sertoli cells.

**Figure 8.**
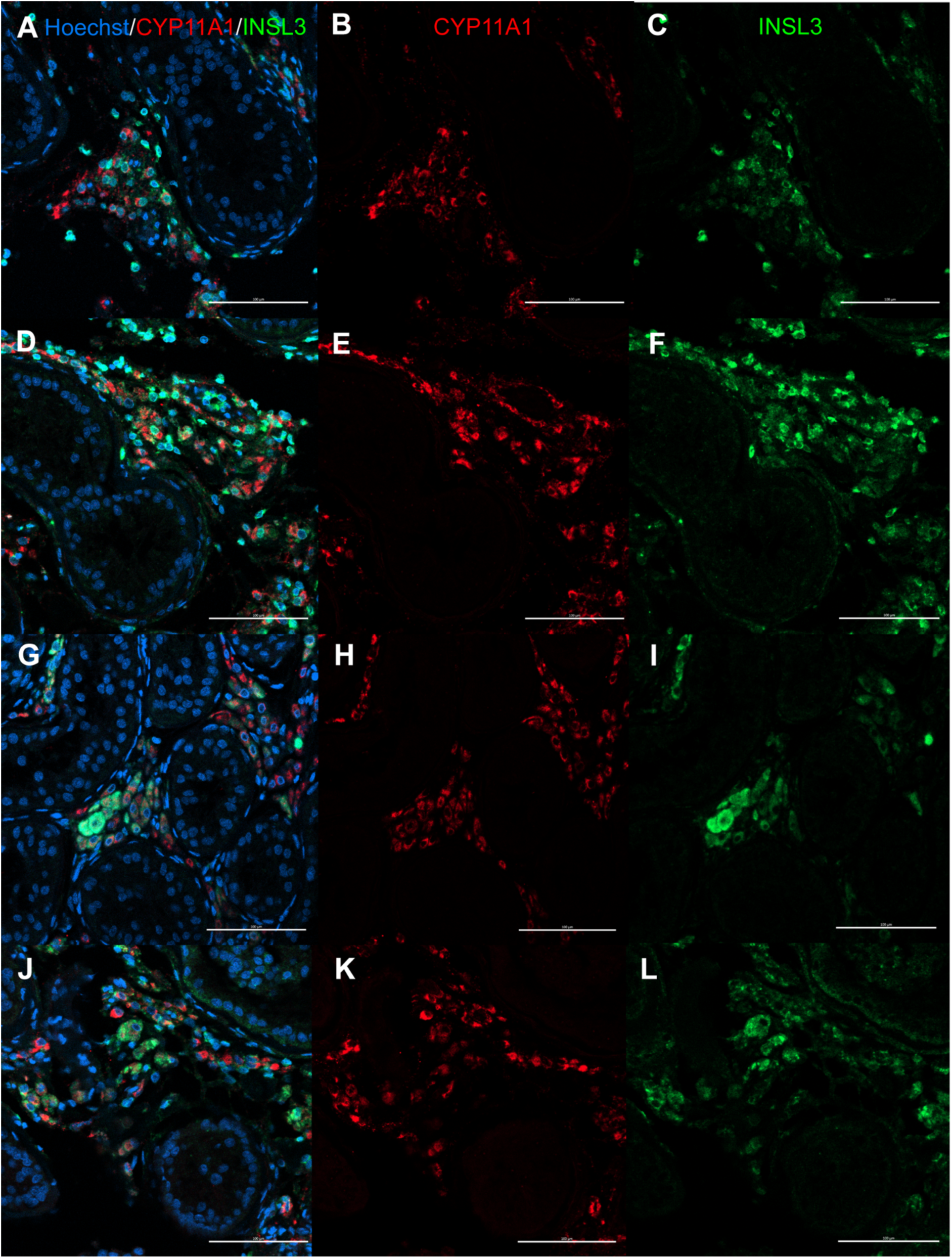
Maturation of Leydig cells. A-C: Tubules with spermatogenesis in intra-testicular graft T4. D-F: Tubules without spermatogenesis in intra-testicular graft T4. G-I: Subcutaneous scrotal graft S2. J-L: Endogenous testicular parenchyma. The majority of Leydig cells co-express CYP11a1 and INSL3. A,D,G,J: Merged pictures. B,E,H,K: CYP11a1 expression for steroidogenic Leydig cells. C,F,I,L: INSL3 expression for mature Leydig cells.

**Figure 9.**
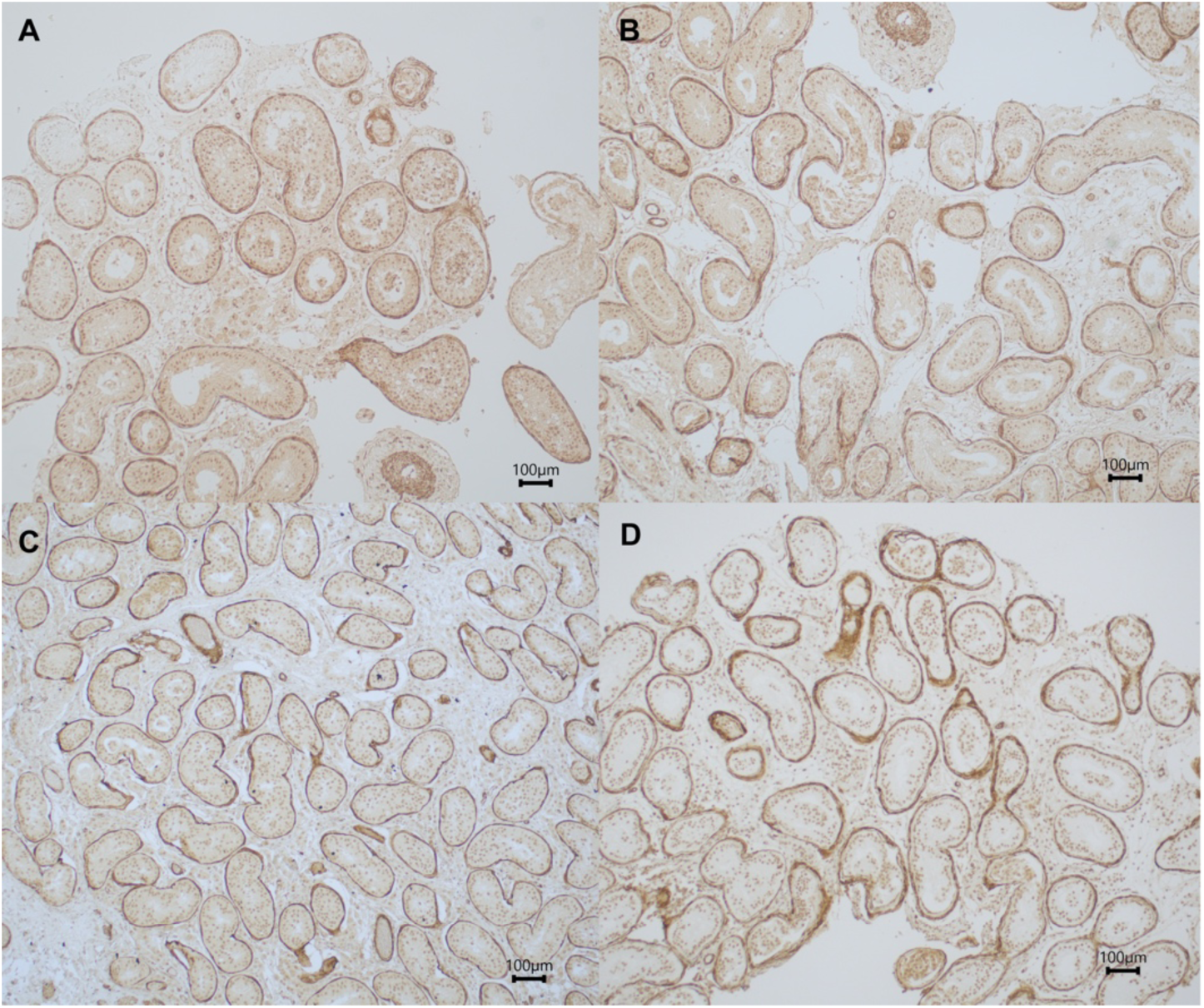
ACTA2 expression in peritubular myoid cells. A: Intra-testicular graft T2 showing tubules with spermatogenesis. B: Intra-testicular graft T2 showing tubules without spermatogenesis C: Subcutaneous scrotal graft S2; D: Endogenous testicular parenchyma. A continuous layer of ACTA2^+^ peritubular myoid cells is observed.

**Table 5.** Somatic cell maturation one year post-transplantation.

| Transplant location | Sertoli cells |  |  | Leydig cells |  | Peritubular myoid cells |
| --- | --- | --- | --- | --- | --- | --- |
|  | SOX9 | AMH | AR | INSL3 | CYP11a1 | ACTA2 |
| T1 | positive | positive | positive | positive | positive | intact |
| T2 | positive | positive | positive | positive | positive | intact |
| T3 | positive | positive | positive | positive | positive | intact |
| T4 | positive | positive | positive | positive | positive | intact |
| S1 | positive | positive | positive | positive | positive | intact |
| S2 | positive | positive | positive | positive | positive | intact |
| S3 | positive | positive | positive | positive | positive | intact |
| S4 | positive | positive | positive | positive | positive | intact |
| TC | positive | positive | positive | positive | positive | intact |
| T1<sub>end</sub> | positive | positive | positive | positive | positive | intact/interrupted |
| T2<sub>end</sub> | ND | ND | ND | ND | ND | ND |
| T3<sub>end</sub> | positive | positive | positive | positive | positive | intact |
| T4<sub>end</sub> | positive | positive | positive | positive | positive | intact |

T: transplanted to the testis; S: transplanted subcutaneously in the scrotum; SOX9: SRY-related HMG-box gene 9 (general marker for Sertoli cells); AMH: anti-Müllerian hormone (marker for immature Sertoli cells); AR: androgen receptor (marker for mature Sertoli cells); INSL3: insulin-like 3 (marker for mature Leydig cells); CYP11a1: cytochrome P450 family 11 subfamily A member 1 (marker for functional Leydig cells); ACTA2: of $\alpha$ -smooth muscle actin (marker for peritubular myoid cells).

Most Leydig cells showed co-expression of INSL3 and CYP11A1, consistent with a mature, steroidogenically active phenotype. However, regions containing Sertoli cell-only tubules in grafts T1 and T4 and adjacent endogenous tissue also harboured INSL3^+^ Leydig cells that lacked CYP11A1, indicating the presence of mature Leydig cells that were not producing testosterone. Conversely, some CYP11A1^+^ Leydig cells lacking INSL3 were also detected (in scrotal grafts and TC, but also in areas with active spermatogenesis in graft T4).

## DISCUSSION

This study presents the first successful autologous transplantation of immature human testicular tissue. Grafted testicular tissue survived, preserved normal architecture and initiated complete spermatogenesis. However, spermatogenesis was identified exclusively in the intra-testicular grafts. Following mechanical mincing and enzymatic digestion, spermatozoa were recovered in the T4 sample, whereas histological examination demonstrated spermatogenic progression in both T2 and T4 grafts. It is important to note that the tissue processed for histology was distinct from the fragment used for enzymatic digestion, which may explain the observed differences between the samples. Histology at location T4 also revealed active spermatogenesis within the endogenous testicular parenchyma. This finding is consistent with the presence of spermatozoa detected by enzymatic digestion at the time of transplantation, indicating that the endogenous tissue already contained focal spermatogenesis prior to grafting. For T2, an endogenous tissue counterpart could not be evaluated at the time of the removal of the grafts one year post-transplantation.

In order not to compromise the feasibility of the transplantation procedure, it is not advisable to wait for the outcomes of the sperm search before starting the thawing process of the cryopreserved immature testicular tissue. Therefore, the decision was taken to proceed with the thawing and transplantation immediately after micro-TESE. Although spermatozoa were found after enzymatic digestion, these were abnormal. Also, spermatozoa that would have been found in the endogenous parenchyma one year post-transplantation would most probably have derived from SSCs exposed to a high-dose of gonadotoxic treatment, with possible negative effects on subsequent fertilization, embryo development, and pregnancy outcome. Notably, all grafts containing spermatogonia also exhibited ongoing spermatogenesis. It remains unclear whether spermatogonia were already absent in the other grafts at the time of transplantation or were lost following transplantation.

The somatic compartment in all evaluated samples exhibited clear signs of maturation, but also maintained features of immaturity, most notably the weak expression of AMH in Sertoli cells. This may indicate that maturation was still in progress. Although most of the Leydig cells showed a mature and steroidogenically active phenotype, some mature Leydig cells within intra-testicular grafts and adjacent endogenous tissue were not producing steroids, maybe due to insufficient LH stimulation. Another notable finding was the presence of CYP11A1⁺INSL3⁻ Leydig cells in specific areas of scrotal grafts and endogenous tissue, but also in tubules with active spermatogenesis (graft T4), suggesting a population of incompletely matured yet steroidogenic Leydig cells (Lottrup *et al*., 2014).

The establishment of complete spermatogenesis after 16 years of cryostorage demonstrates that the applied cryopreservation protocol using 1·5 M DMSO combined with 0·15 M sucrose and 10% HSA (Baert *et al*., 2018) effectively preserves the viability and maturational capacity of both somatic and germ cells. The survival of all grafts further supports the robustness of this approach. These findings contrast with observations in a rat model, in which testicular tissue stored for 20 years exhibited impaired progression of spermatogenesis (Whelan *et al*., 2022). Notably, their cryopreservation medium did not contain sucrose. The addition of sucrose has previously been shown to enhance spermatogonial survival during cryopreservation (Baert *et al*., 2013), which may have contributed to the preserved developmental potential observed in our grafts.

Endocrine and semen parameters remained unchanged during the one-year follow-up period after transplantation. This was expected, as the transplanted fragments represent only a very small proportion of the total testicular volume, making measurable alterations in circulating FSH, LH, testosterone, or INHB concentrations unlikely, even though vascularisation of the grafts was successfully established. Similarly, no changes in semen characteristics were anticipated. The seminiferous tubules within the scrotal, but also the intra-testicular, grafts do not make connections with the rete testis, preventing spermatozoa produced within the grafts from entering the ductal system. Therefore, to enable future attempts at achieving biological parenthood, retrieval of the grafts or micro-TESE in combination with ICSI after testicular tissue grafting will be necessary.

A one-year interval between transplantation and graft retrieval appears appropriate, as complete spermatogenesis was established by this time. Earlier retrieval might not have allowed maturation of the somatic niche or completion of germ cell differentiation, as was observed in human-to-mouse grafting experiments (Van Saen *et al*., 2011, 2013). Whether prolonged retention of grafts *in situ* could eventually lead to degeneration remains uncertain, particularly because spermatozoa produced within the grafts cannot exit the tissue. Evidence from animal studies suggests that grafts may have a limited functional lifespan (Abrishami *et al*., 2010; Gourdon and Travis, 2011).

Fixation of the grafts was performed using both Prolene and silk sutures. Both materials remained identifiable, but it was easier to identify the Prolene sutures than the silk sutures at the time of graft retrieval. Prolene sutures, however, were not detectable by ultrasound and demonstrated less vascularization. Silk and Prolene interact with human tissue differently, affecting neo-angiogenesis. Silk, a natural protein, causes more inflammation than synthetic materials (Alves de Oliveira et al., 2024). While inflammation is necessary for healing, too much can hinder grafts. Neo-angiogenesis is part of tissue healing. Studies show that silk supports endothelial cell growth and angiogenesis when used as a scaffold (Li et al., 2024).

Due to the use of non-resorbable sutures, the grafts could be easily located. Transplanted tissue fragments were clearly distinguishable from the surrounding endogenous parenchyma. Whereas the grafts exhibited a firmer and more consolidated architecture, endogenous tissue appeared softer and less compact. At retrieval, each graft had increased substantially in size. However, during follow-up, the growth was not detectable by ultrasound. Small tissue pieces that were transplanted together (location T4 and S4) could, on histological sections, still be identified as separate fragments next to each other. Notably, grafts placed under the scrotal skin displayed extensive areas of fibrosis, which might be due to the fact that the scrotal grafts experienced a longer ischaemic period. Indeed, no vascularization was seen in scrotal grafts at the first follow-up visit. Whether extensive fibrosis was also observed in animal studies, has not been reported.

Interpretation of differences between intra-testicular and subcutaneous scrotal grafts must be approached with caution, given the focal and extremely low number of spermatogonia present in the cryopreserved tissue. Although intra-testicular grafts demonstrated spermatogenic activity and less fibrosis, at this moment, it is too early to draw firm conclusions on optimal grafting site, optimal fragment size or the most suitable suture material for graft fixation. These variables require systematic evaluation in future studies. Direct comparisons between intra-testicular and subcutaneous scrotal grafts have not yet been investigated in animal models. Existing studies have instead compared intra-testicular grafts with grafts transplanted under the dorsal skin (Ntemou *et al*., 2019; Hutka *et al*., 2020a) or subcutaneous grafts placed in the scrotum to those positioned under the dorsal skin (Fayomi *et al*., 2019). Xenograft studies demonstrate that graft location substantially influences germ cell survival and development. Intra-testicular xenotransplantation supported the differentiation of immature germ cells from marmoset (*Callithrix jacchus*) into spermatozoa, while in subcutaneous transplants, spermatogenic arrest was observed (Ntemou *et al*., 2019). In human-to-mouse xenograft studies, germ cell survival was higher in intra-testicular grafts compared to subcutaneous grafts (Hutka *et al*., 2020a). However, autologous transplantation of immature non-human primate tissue to locations under the dorsal and scrotal skin has been shown to support spermatogenesis at both sites (Fayomi et al., 2019). Research into comparing different suture materials for grafting in animal models has not yet been carried out.

Since pre-transplant gonadotrophin levels were already high (similar or even higher than levels observed in pubertal boys between 13-17 years: 9·8 IU/l for LH; 12·9 IU/l for FSH), exogenous hormonal stimulation does not appear to be required to initiate spermatogenesis in intra-testicular grafts. This is consistent with previous work demonstrating that exogenous administration of FSH did not influence meiotic differentiation in human testicular xenografts (Van Saen *et al*., 2013). Whether hormonal supplementation might benefit scrotal grafts remains uncertain. It is possible that exogenous gonadotrophins enhance graft efficiency by promoting Sertoli and Leydig cell maturation, as suggested in a study in which foetal human tissue was xenografted to castrated mice (Hutka *et al*., 2020b).

This study has some important limitations. Only a single patient was included and the cryopreserved tissue contained an extremely low number of SSCs, which limits the generalisability of the findings. We noted that the reliability of non-invasive parameters, such as ultrasonography, serum hormone levels, and semen analyses, is limited in predicting graft growth and ongoing sperm production within grafts. More accurate biomarkers and refined monitoring strategies are needed. Moreover, the fertilisation and developmental potential of the spermatozoa recovered from the graft remains unknown. Future research will require larger cohorts and systematic evaluation of graft performance across different transplantation sites and tissue sizes. As the patient intends to pursue biological parenthood, close monitoring of embryo development, pregnancy progression, and long-term health outcomes in any resulting offspring will be essential to ensure both safety and efficacy of this fertility restoration approach.

To conclude, this study demonstrates that complete spermatogenesis can be achieved more than sixteen years after tissue storage, despite the exceptionally low number of SSCs present in the cryopreserved tissue. Our observations suggest that intra-testicular grafting may offer advantages over subcutaneous placement. However, this finding requires confirmation in additional studies.

## DATA AVAILABILITY

Data obtained from histological, immunohistochemical and immunofluorescent analyses are available at https://doi.org/10.5281/zenodo.18739648.

The dataset supporting the clinical outcomes of this study is securely stored in the Vrije Universiteit Brussel (VUB) Institutional Data Repository under restricted access, with the accession number VUB/GRAD/1/000021, to protect participant privacy. Access requests will be reviewed on an individual basis and must be directed to Prof. Ellen Goossens. She will assess the request, considering the research purpose and potential commercial applications. Prior to any data sharing or release, a data use agreement, in compliance with the VUB legal department’s guidelines, must be completed and signed.

Metadata for the datasets is available through the VUB Research Portal at: https://researchportal.vub.be/en/datasets/first-in-human-transplant-of-immature-testicular-tissue-for-ferti/ and https://researchportal.vub.be/en/datasets/first-in-human-transplant-of-immature-testicular-tissue-for-ferti-2/

## Data Availability

Data obtained from histological, immunohistochemical and immunofluorescent analyses are available online at https://doi.org/10.5281/zenodo.18739648.
Clinical data produced in the present study is available upon reasonable request to the authors.
Metadata for the datasets is available through the VUB Research Portal at: https://researchportal.vub.be/en/datasets/first-in-human-transplant-of-immature-testicular-tissue-for-ferti/ and https://researchportal.vub.be/en/datasets/first-in-human-transplant-of-immature-testicular-tissue-for-ferti-2/

## ACKNOWLEDGEMENTS

The authors would like to thank Dr. Segers (embryologist Brussels IVF) for assisting with preparing the testicular tissue prior to transplantation, Mrs. Nulens and Mrs. Illingworth (study nurses Brussels IVF) for planning screening and follow-up tests, the entire team of operating theatre nurses, Dr. Uvin for assisting during surgery, Mrs. Wieme for semen analyses, and to the Clinical chemistry and Radioimmunology laboratory for blood analyses.

## AUTHORS’ ROLES

All authors should confirm that they had full access to all the data in the study and accept responsibility to submit for publication. E.G. contributed to study conception and design, funding acquisition, supervision of the study, data collection, interpretation of the results, and writing the original draft of the manuscript. V.V. contributed to study conception and design, performed the consultations and the actual transplantation procedure, and contributed to validation of the results, review and editing of the manuscript. E.D.B and E.D. performed the histological evaluation, and contributed to the validation of the results, review and editing of the manuscript. I.M. thawed and prepared the testicular tissue prior to transplantation, and reviewed the manuscript. C.E. performed ultrasound and colour-Doppler, and reviewed the manuscript. W.W for conducting the histopathological evaluation, and review and editing of the manuscript. I.G. contributed to blood analyses, funding acquisition, and review and editing of the manuscript. H.T. contributed to study conception and design, funding acquisition, supervision of the research and validation of the results, and review and editing of the manuscript.

